# Near-infrared II hyperspectral imaging improves the accuracy of pathological sampling of multiple cancer specimens

**DOI:** 10.1101/2022.10.27.22281545

**Authors:** Lingling Zhang, Jun Liao, Han Wang, Meng Zhang, Dandan Han, Chen Jiang, Zhanli Jia, Yao Liu, Chenchen Qin, ShuYao Niu, Hong Bu, Jianhua Yao, Yueping Liu

## Abstract

Pathological histology is the clinical gold standard for cancer diagnosis. Incomplete or excessive sampling of the formalin-fixed excised cancer specimen will result in inaccurate histology assessment or excessive workload. Conventionally, pathologists perform specimen sampling relying on naked-eye observation which is subjective and limited by human perception. Precise identification of tumor beds, size, and margin is challenging, especially for lesions with inconspicuous tumor beds. To break the limits of human eye perception (visible: 400-700 nm) and improve the sampling efficiency, in this study, we propose using a second near-infrared window (NIR-II: 900-1700 nm) hyperspectral imaging (HSI) system to assist specimen sampling on the strength of the verified deep anatomical penetration and low scattering characteristics of the NIR-II optical window. We use selected NIR-II HSI narrow bands to synthesize color images for human eye observation and also apply artificial intelligence (AI)-based algorithm on the complete NIR-II HSI data for automatic tissue classification to assist doctors in specimen sampling. Our study employing 5 pathologists, 92 samples and 7 cancer types shows that NIR-II HSI-assisted methods have significant improvements in determining tumor beds compared with conventional methods (Conventional color image with or without X-ray). The proposed system can be easily integrated into the current workflow, and has high imaging efficiency and no ionizing radiation. It may also find applications in intraoperative detection of residual lesions and identification of different tissues.

## Introduction

After excised cancer tissues are fixed in formalin, pathologists manually perform gross visual examination and specimen sampling. Precise identification of tumor beds, size, and margin is challenging due to the limited visual contrast between cancer and its adjacent tissues. Hence, there is a great need for an auxiliary tool to help pathologists to improve their work quality and efficiency.

Researchers are exploring ways to help identify the area of the tumor bed; among these, X-rays, a more penetrating form of radiation, have been used in guiding pathologists in the sampling of breast specimens^1^. However, X-rays provide image contrast relying on density differences, therefore are not always effective in differentiating fibers and cancerous foci which are both dense tissues; moreover, the presence of ionizing radiation and the high cost of X-rays make this methodology unpopular in pathology sampling departments. In addition to X-rays, new techniques for the detection of intraoperative cancer margins have emerged in recent years that are worthy of use as tools for sampling. One such technique is the application of fluorescence multi-spectroscopy to locate liver cancers in vivo for precision surgery^2^. The MarginProbe is an instrument developed for intraoperative detection of breast cancer margins based on radiofrequency spectroscopy technology^3^. In addition, academic studies have proposed the use of various advanced optical techniques for intraoperative cancer margin detection including terahertz^4,5^, ultrasound^6–8^, and optical coherence tomography (OCT)^9–12^. These methods either require long image acquisition time or trained professional technicians, hence may slow down or add significant cost to the specimen sampling workflow. Hyperspectral imaging (HSI), a spectroscopy-based imaging method which collects hundreds of images at different wavelengths for the same spatial area, helped in the evaluation of resection margins of intraoperatively resected fresh breast tissue specimens^13–17^ and hyperspectral non-contact endoscopic diagnostic system imaging in vivo^18,19^ helped determine the benignancy/malignancy of colon cancers and detect early malignant lesions. Since comprehending dozens or hundreds of hyperspectral images is time-consuming and difficult, most of these hyperspectral methods are limited to visible to NIR-I (400-900 nm) and only provide the final artificial intelligence (AI)-based segmentation results without intermediate images to which physicians can refer as the reference. Moreover, most of these data are based on in vivo or fresh ex vivo tissue studies and the ability to discriminate formalin-fixed cancer tissues remains unknown^20^.

Inspired by Hong’s and Hu’s work which showed excellent anatomical penetration and low scattering ability of the NIR-II optical window on in vivo fluorescence imaging of mouse hind limb and brain and human lung^21,22,2^, we believe, together with hyperspectral imaging technique which produces rich spectral and spatial information of the specimen, NIR-II bright field imaging can be introduced to pathology workflow as well.

This study is the first report on the analysis of the ability of NIR-II HSI to assess postoperative formalin-fixed cancer tissues from different systems. We investigated the spectral absorption differences at 900–1700 nm for each tissue type and selected characteristic bands to generate information-rich NIR-II synthesized color images for direct observation by physicians. We also performed spectral analysis of the entire HSI using an AI algorithm to provide intelligent and automatic segmentation of different tissue regions to assist pathologists in identifying cancer tissues. Through the validation of 62 cancer tissue specimens, our method with the combination of the generated NIR-II synthesized color images and AI segmentation results showed a higher cancer detection accuracy compared to that of the conventional methods.

## Results

### 1. The NIR-II hyperspectral imaging system and representative results

A NIR-II HSI system (Gaiafield N17E by Dualix Spectral Imaging, resolution: 320*256) was used to obtain NIR-II hyperspectral images with wavelengths from 900-1700 nm. The imaging system setup is shown in (Fig. 1a). A tissue sample was first placed on a stage that could be panned left and right and a linear light source derived from a halogen lamp was used for illumination. After the sample was illuminated by the linear light source, the broad-spectrum light interacted with the tissue through absorption and reflection. The reflected light was captured by a NIR-II hyperspectral camera and the broad-spectrum signal was divided into a total of 256 narrow bands from 900–1700 nm with a spectral resolution of 3.2 nm by the spectroscopic device inside the hyperspectral camera. A three-dimensional data cube (Fig. 1b) was obtained via horizontal scanning of the stage. After whiteboard reflectance correction, the reflectance spectral curves of different regions of the tissue were obtained (Fig. 1c). In Fig. 1d, we show the typical NIR-II hyperspectral images of a human colon specimen using the proposed system. Instead of displaying all the 256 bands, here, we show three selected characteristic bands: 1100 nm, 1300 nm and 1450 nm. We also composite a NIR-II color image using the three bands. The NIR-II synthesized color image is closer to the human eye’s viewing habits and is more informative than monochrome images, allowing physicians to identify different tissues. Compared with the conventional color image, the NIR-II synthesized color image demonstrates better contrast for cancer, normal mucosa, muscle and adipose tissue as shown in Fig. 1d, the cancer tissue (red box indicated) appears white-orange, the normal mucosa (green box indicated) appears slightly darker than the cancer tissue, the muscle tissue (purple box indicated) appears darker orange than the cancer tissue, and the adipose tissue (yellow box indicated) appears bright yellow. However, in the conventional color image, cancer, normal and muscle all looks white and have close intensity and low contrast. Pathologists can identify the cancer area with more confidence with the NIR-II synthesized image compared to naked eye observation.

**Fig.1.**
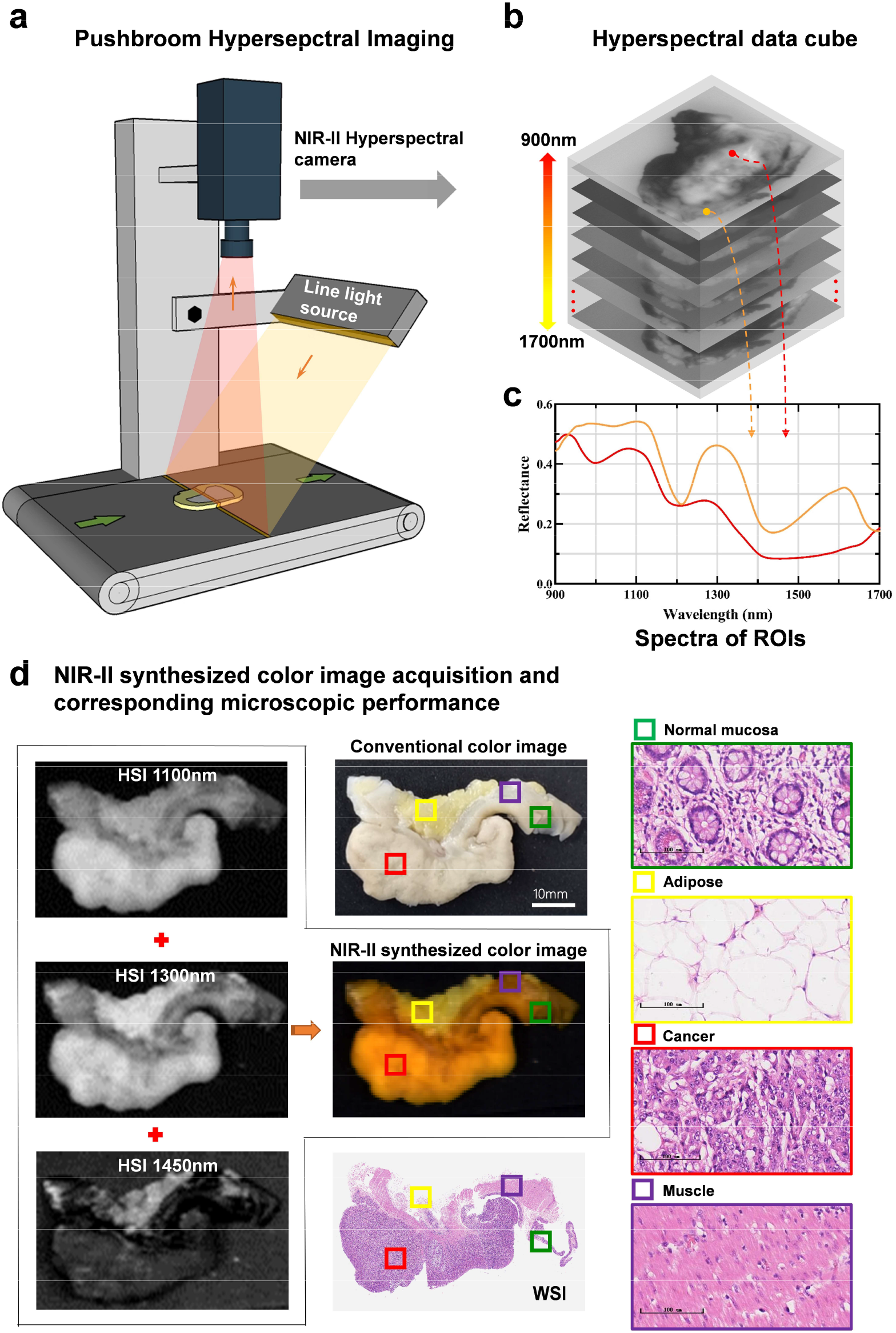
Schematic diagram of NIR-II hyperspectral imaging system and typical spectral and narrowband spatial data. **a** Tissue samples were placed on a translation table, and the 3D hyperspectral data cube was obtained by push-scan imaging. **b** Each hyperspectral data cube contains NIR-II spectral and spatial information ranging from 900 nm to 1700 nm. **c** After whiteboard reflectance correction, two spectral curves for regions of interest in b. **d** Typical hyperspectral images at 1100 nm, 1300 nm and 1450 nm for a colon specimen. We also show the NIR-II synthesized color image using these three narrow bands and corresponding tissue appearance under microscope. The red boxes indicate areas of cancerous tissue, the yellow boxes indicate adipose tissue, the green boxes indicate normal mucosa, and the purple boxes indicate muscle tissue. Microscopic (HE ×200).

### 2. Preparation of tissue samples and experimental procedures

As shown in Fig. 2a, the surgically excised specimens were placed in a sufficient amount of 10% neutral formalin solution within 30 minutes after isolation and then fixed for 12–48 h. The excised tissue pieces were 5mm±1mm (average 5mm) and contained cancer tissues with 1–2 cm of surrounding normal tissues. A total of 92 samples included 63 from hollow organ tissues (17 cases of colon cancer, 18 cases of rectal cancer, 9 cases of esophageal cancer, and 19 cases of gastric cancer) and 29 from parenchymal organ tissues (12 cases of lung cancer, 8 cases of kidney cancer, and 9 cases of breast cancer) were collected for this study. All specimens had conventional color images taken from the same viewing angle as the hyperspectral images, as well as the X-ray images taken using a cabinet X-ray imaging system (Fig. 2b). To obtain gold standard of tissue identification, the tissue slices were then routinely dehydrated, embedded in paraffin, stained with hematoxylin and eosin (HE), and scanned to obtain whole-slide images (WSI). The WSI fragments were scanned and virtually stitched together using an in-house developed WSI-stitching software to restore virtual large sections (Fig. 2c). 30 cases were used to train the AI algorithm and train doctors on how to read images for annotation. 62 cases were used for test verification. The pathologists used the ASAP^23^ annotation software to perform pathological annotation on the virtual large section and used the results as the ground truth. The hollow organs were labeled as cancer (red), normal mucosa (green), adipose (yellow), or muscle tissue (purple). The parenchymal organs were labeled as cancer (red), normal (green), or adipose tissue (yellow). Finally, the breast tissues were labeled as cancer (red), adipose (yellow), or fibrous connective tissue (green).

**Fig.2.**
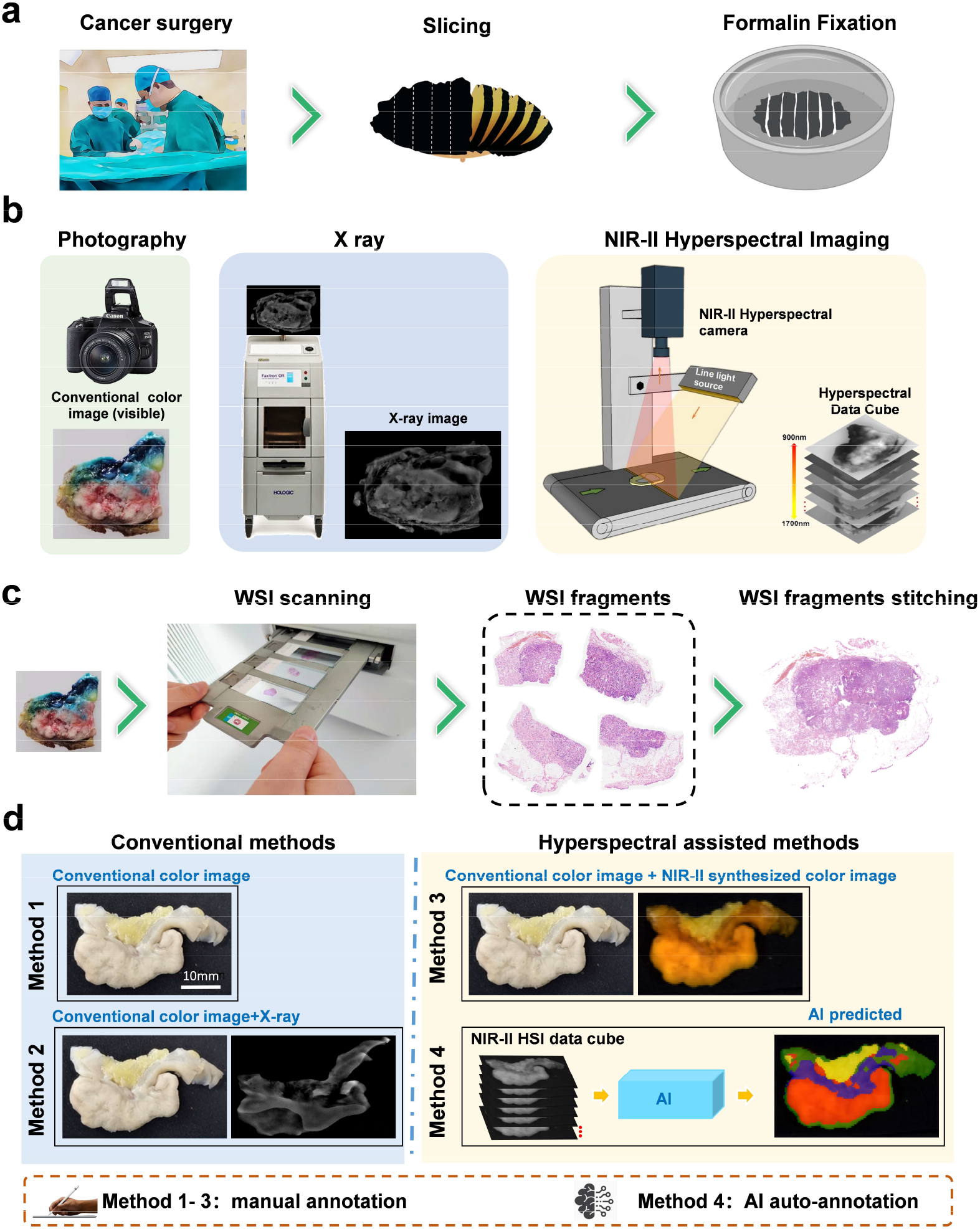
Experimental flow chart. **a** In vitro specimens were cut open and fixed in 10% formalin solution. **b** Conventional color images (400–700 nm), X-ray images and NIR-II hyperspectral images were obtained respectively. **c** Tissue samples were prepared by conventional dehydrated paraffin embedding, HE stained sections were scanned using WSI scanner and digitally stitched together to restore the virtual large slice using an in-house developed WSI stitching software. **d** Pathologists labeled the cancer area with different evidence (Method 1-3) with two-week forgetting periods. Method 4 is an auto-segmentation method (AI prediction). Method 1: Referring to conventional color image only. Method 2: Referring to X-ray image and conventional color image. Method 3: Referring to NIR-II synthesized color image and conventional color image. Method 4: AI prediction results without manual annotation.

### 3. Hyperspectral image processing

The acquired hyperspectral images underwent reflectance correction (see methods) first and then two images were obtained and used as physician sampling references. The first image was a NIR-II synthesized color image in selected optimal bands (1100 nm for red channel, 1300 nm for green channel, and 1450 nm for blue channel) according to the clustering-based band selection algorithm and pathologists’ manual selection^24^. The second image was the result of tissue classification using a support vector machine (SVM)^25,26^, a supervised machine learning algorithm, by taking all the 256 collected bands into the spectral analysis and classification. The training data and the test data we use for the SVM are shown in Table.1.

**Table 1.**
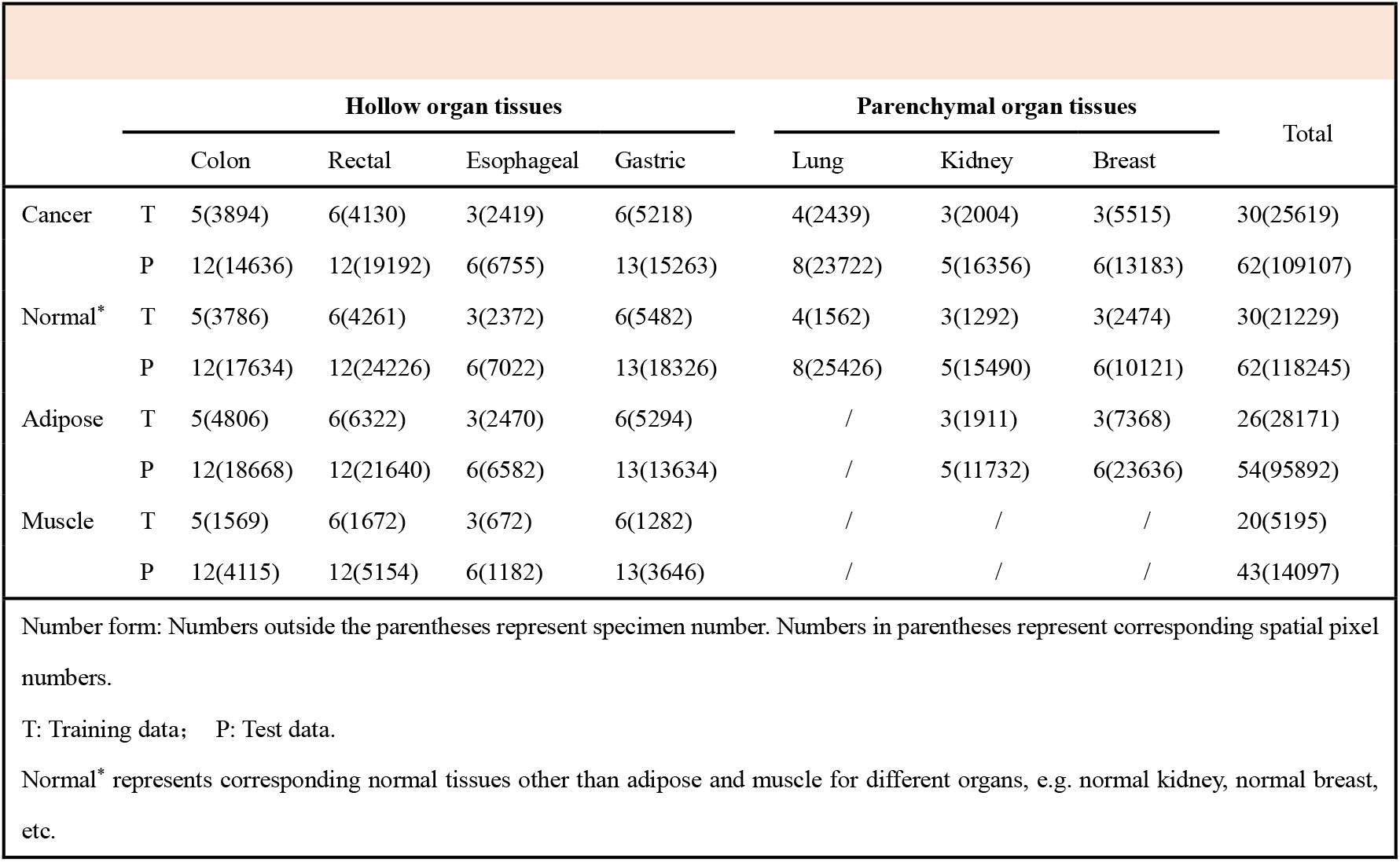
Data Description.

| Table 1 Data Description |  |  |  |  |  |  |  |  |  |
| --- | --- | --- | --- | --- | --- | --- | --- | --- | --- |
|  |  | Hollow organ tissues |  |  |  | Parenchymal organ tissues |  |  | Total |
|  |  | Colon | Rectal | Esophageal | Gastric | Lung | Kidney | Breast |  |
| Cancer | T | 5(3894) | 6(4130) | 3(2419) | 6(5218) | 4(2439) | 3(2004) | 3(5515) | 30(25619) |
|  | P | 12(14636) | 12(19192) | 6(6755) | 13(15263) | 8(23722) | 5(16356) | 6(13183) | 62(109107) |
| Normal* | T | 5(3786) | 6(4261) | 3(2372) | 6(5482) | 4(1562) | 3(1292) | 3(2474) | 30(21229) |
|  | P | 12(17634) | 12(24226) | 6(7022) | 13(18326) | 8(25426) | 5(15490) | 6(10121) | 62(118245) |
| Adipose | T | 5(4806) | 6(6322) | 3(2470) | 6(5294) | / | 3(1911) | 3(7368) | 26(28171) |
|  | P | 12(18668) | 12(21640) | 6(6582) | 13(13634) | / | 5(11732) | 6(23636) | 54(95892) |
| Muscle | T | 5(1569) | 6(1672) | 3(672) | 6(1282) | / | / | / | 20(5195) |
|  | P | 12(4115) | 12(5154) | 6(1182) | 13(3646) | / | / | / | 43(14097) |
| Number form: Numbers outside the parentheses represent specimen number. Numbers in parentheses represent corresponding spatial pixel numbers.<br>T: Training data; P: Test data.<br>Normal* represents corresponding normal tissues other than adipose and muscle for different organs, e.g. normal kidney, normal breast, etc. |  |  |  |  |  |  |  |  |  |

### 4. Experimental verification

The 62 samples of the test set include 37 samples of hollow organ tissue (10 colon cancer, 13 rectal cancer, 4 esophageal cancer, and 10 gastric cancer) and 25 samples of solid organ tissues (11 lung cancer, 7 kidney cancer, and 7 breast cancer). Five pathologists with more than three years of experience marked the areas of cancer with three different methods by referring to different evidence (as shown in Fig. 2d, Methods 1-3). Method 4 represents the AI predicted results and involves no manual annotation as Methods 1-3 do.

The “gold standard” was formulated after the above tests by 5 pathologists referring to WSI and all image data. The WSI was used as a primary reference and combined with all available images (X-rays, NIR-II synthesized color image, AI predicted image) as secondary evidence. The final results discussed by the five doctors were defined as the “gold standard”.

A two-week gap is set as the forgetting period between the annotation of different methods. The WSI, X-ray and HSIs are co-registered (see method) with the conventional color images. After registration, all the annotations were mapped onto the conventional image for statistical evaluation on the same scale.

### 5. The spectral profiles of different cancer species

The spectral profiles of different tissues in different organs we collected are shown in Fig. 3. These curves are calculated by averaging all the collected data according to the gold standard annotation. At around 1300 nm, the adipose has a reflection peak and demonstrates good contrast with other types of tissues. 1100 nm is another peak band helpful to differentiate cancer and normal tissues by comparing the peak difference. All these tissue types have a trough of the spectral curve at 1450nm. We can observe that the adipose and muscle have very similar curve shapes among all collected specimens. The spectral curve of cancer and normal tissues for hollow organs are also similar but different from that of parenchymal organs. For hollow organs, the reflectance of cancer is larger than the normal mucosa, therefore we can observe that the cancer area of the colon specimen in Fig. 1d is brighter than the normal mucosa in the NIR-II synthesized image.

**Fig.3.**
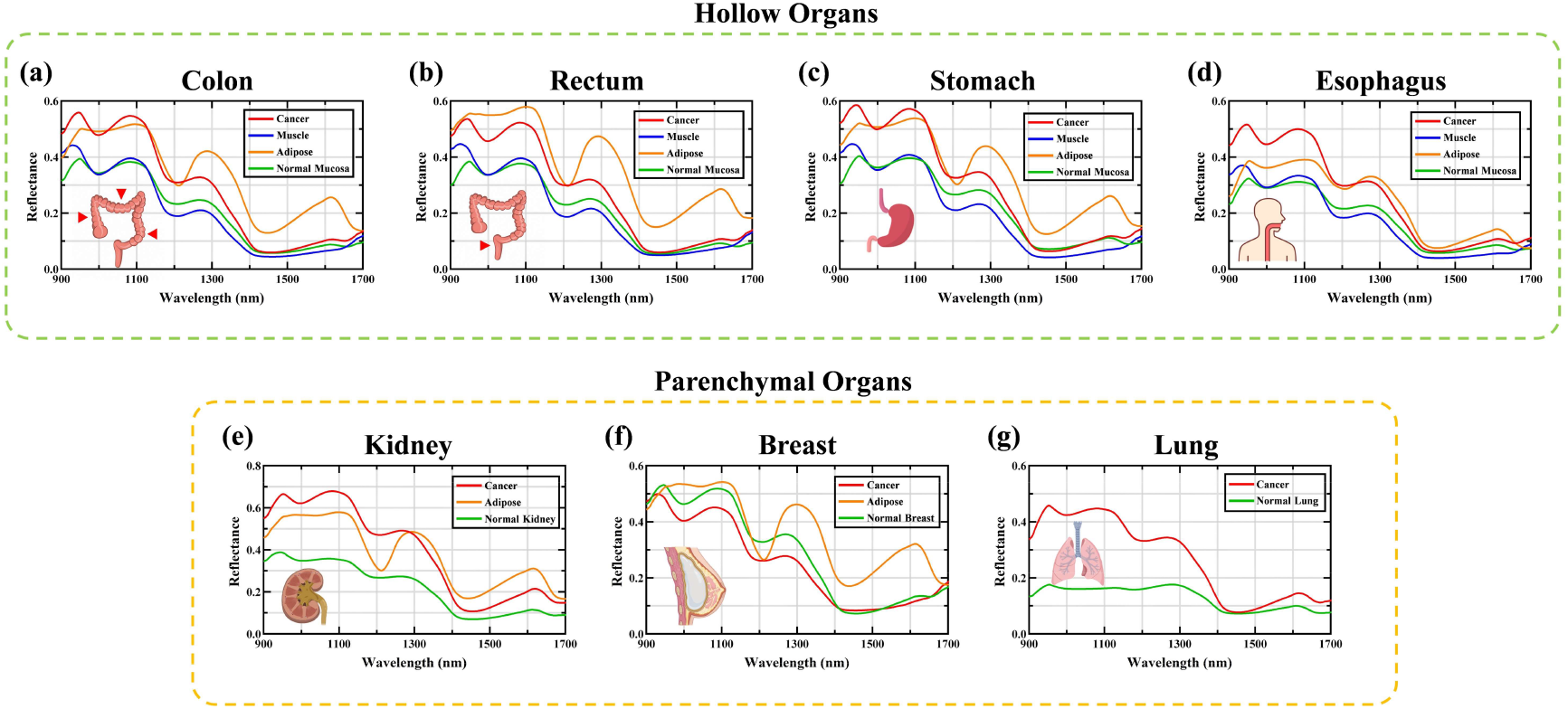
Spectral profiles of different tissue types in different organs.

### 6. Identification of hollow organ cancer with NIR-II HSI

The hollow organ tissues in this study mainly include those from colon, rectal, gastric, and esophageal cancers. In Fig. 2d, we already showed the example for colon. Fig. 4 shows the representative results of rectal cancer, gastric cancer and esophageal cancer. In all these cancer specimens, compared to the conventional color images (Fig. 4a1-3) and X-ray images (Fig. 4b1-3), NIR-II synthesized color images (Fig. 4c1-4) show improvements in determining tissue (adipose, muscle and tumor) margins. From Fig. 4c1-3, the cancer areas are brighter than normal mucosa and muscle, making cancer identification much easier than conventional color images and X-ray. The adipose under NIR-II color looks bright yellow which takes no effort to identify. X-ray imaging is density-sensitive and works well for breast cancer specimens (Fig. 5a1) but falls short for hollow organs. Here, X-ray shows good boundary between adipose and other tissues but fails to distinguish the muscularis structure, normal mucosa and cancer. Our AI predicted classification results provide a pixel-level segmentation by considering all the collected wavebands as shown in (Fig. 4d1-4). Using the stitched histology images in Fig. 4 e1-3 as the gold standard, the AI predicted results match excellently with actual corresponding tissues. It is worth mentioning that to make full use of HSI data, we use all the 256 wavebands for AI training and prediction instead of using selected bands. The training process is a data-driven method for the AI to automatically find the principle contributing wavebands and spectral features.

**Fig.4.**
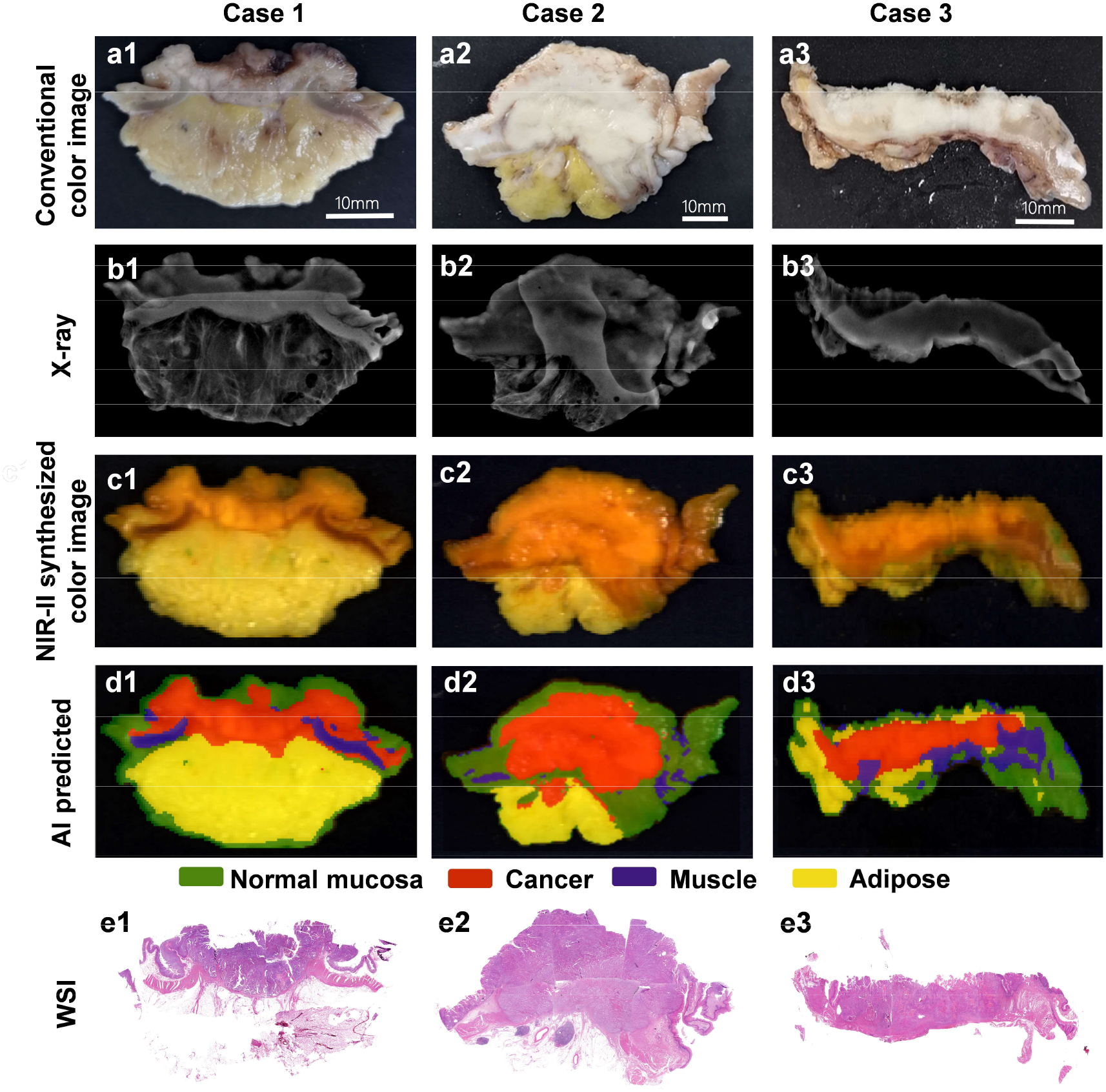
Image classification of different tissues in hollow organs. Case 1: rectal cancer tissue; Case 2: gastric cancer tissue; Case 4: esophageal cancer tissue. **a1-3** Conventional color images of the three specimens. **b1-3** X-ray images. **c1-3** NIR-II synthesized color images. **d1-3** AI predicted segmentation results. Red is cancer tissue, purple is muscularis, green is normal mucosa, yellow is adipose tissue. **e1-3** The stitched WSI of the three specimens.

**Fig.5.**
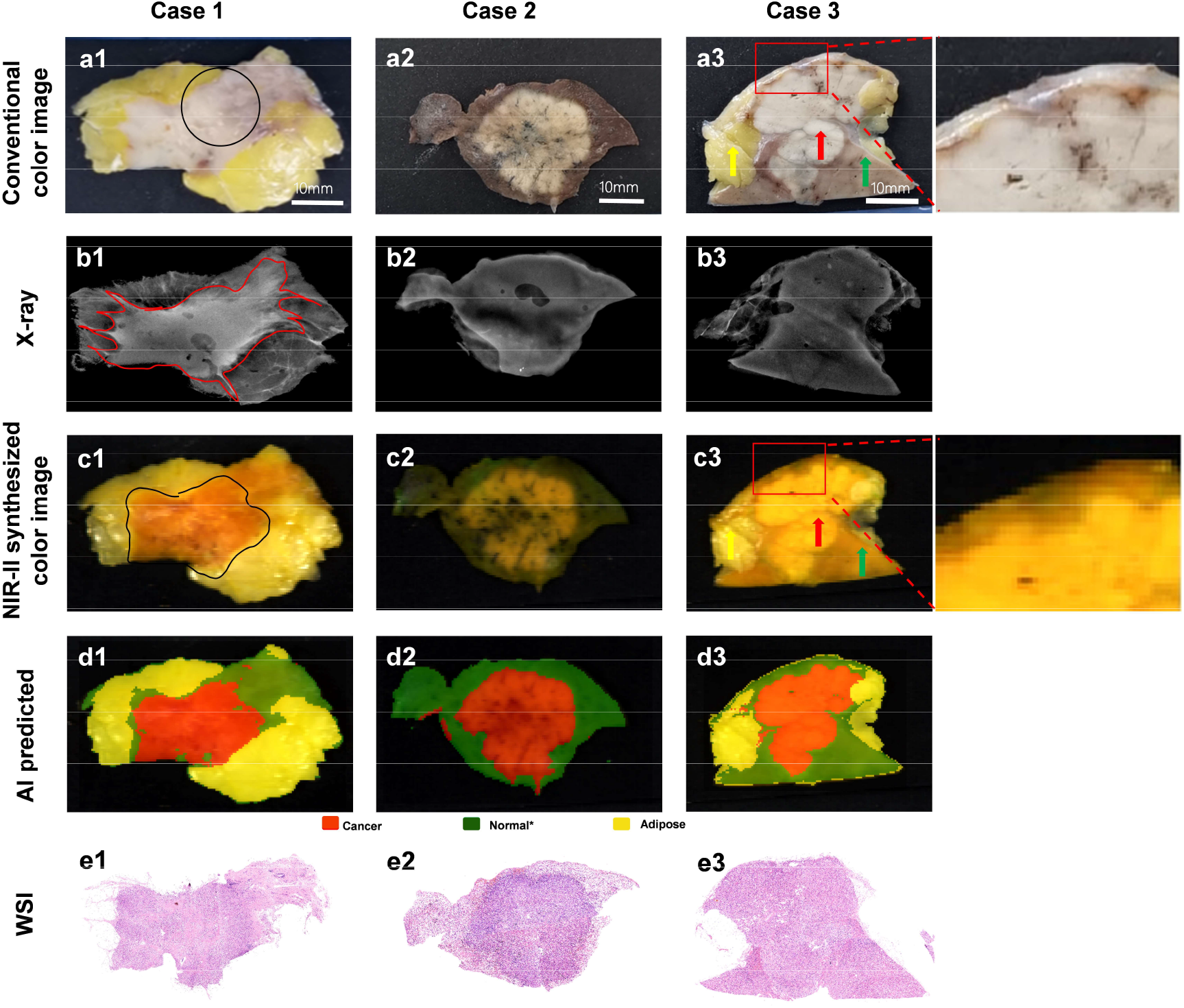
Representative results of parenchymal organ cancer. Case 1: Breast Cancer. Case 2: Lung cancer. Case 3: Kidney Cancer. **a1** The black circle indicates a boundary between cancer and normal breast tissues in the conventional color images. **b1** X-ray images with the dense tissue marked by red lines. **c1** The NIR-II synthesized color image with the cancer tissue marked by black lines. **d1-3** AI predicted results. ^*^ Normal stands for normal breast, normal lung and normal kidney. **e1-3** The stitched WSIs. **a2** Conventional color image of lung cancer. **b2** The X-ray image fails to show the boundary of the tumor. **c2** The NIR-II synthesized color image with the cancer tissue grayish-yellow. **a3** Conventional color image of kidney cancer and local magnification of cancer boundary. **b3** X-ray image of kidney fails to show tissue boundaries. **c3** NIR-II synthesized color image and local magnification of cancer boundary.

### 7. Identification of parenchymal organ cancer tissue with NIR-II HSI

The parenchymal organ cancer tissues we collected for this experiment were kidney, lung, and breast cancers. In the conventional color image of breast cancer (Fig. 5a1), we can vaguely find cancer and normal breast boundary indicated by the black circle with naked eye. In (Fig. 5b2), X-rays show good contrast between low-density tissue(fat) and high-density tissue (cancer and normal breast) but fail to differentiate cancer and normal boundary as the red box indicated. In the NIR-II synthesized color image, the adipose tissue is bright yellow (Fig. 5c1). The cancer area shows a white-orange mixed color compared to the neighboring normal breast tissue which has a similar but more transparent-like color. The NIR-II color image which is composited with 3 selected HSI bands still has difficulty in providing better contrast of cancer and normal breast than conventional color image. However, in the AI-predicted image (Fig. 5d1), the red area shows the cancer contour closest to the one shown on WSI (Fig. 5e1). The AI algorithm which analyzes the entire hyperspectral data cube can provide a clearer boundary between cancer and normal breast area than all the other image evidences.

In lung cancer samples, our experimental results show that HSI can help distinguish tumor tissue from normal lung tissue with good contrast as well. In the NIR-II synthesized color image, the tumor tissue appears grayish yellow (Fig. 5c2). The AI segmentation image can accurately predict the tumor tissue as well (Fig. 5d2).

For kidney specimen, as shown in (Fig. 5a3), conventional color images of the cancer tissue appear grayish-white, while adipose and normal kidney tissues appear yellow and light brown, respectively; however, the zoomed image shows that the border between cancer and normal kidney tissues is not easily distinguishable. The X-ray image shows worse cancer boundaries (Fig. 5b3). In the NIR-II synthesized color image (Fig. 5c3), the kidney cancer tissue appears white-orange (red arrow pointed), the adipose tissue appears bright yellow (yellow arrow pointed), and normal kidney tissue appears darker orange-yellow (green arrow pointed). The magnified image shows a distinguishable boundary between cancer and surrounding normal kidney tissues. In the AI-predicted results, the cancer outline is more consistent with the cancer boundary in WSI (Fig. 5e3). For kidney tumors, according to the AJCC TNM staging criteria^27^, if the cancer is confined to the kidney, the T staging is T1-T2. Once it breaks through the renal capsule and invades the surrounding adipose tissue, the T staging should rise to T3. Therefore, finding the location of the renal capsule invasion is crucial during the sampling process, since it is directly related to the clinical stage of the patient and affects the subsequent diagnosis, treatment plan, and prognosis. In Case 3, the conventional color image in Fig. 5a3 shows suspicious renal capsule invasion in the zoomed image. However, the zoomed area of NIR-II synthesized color image (Fig. 5c3) shows a clearer cancer boundary and no renal capsule invasion in this location with confidence. This helps achieve accurate sampling and avoid unnecessary extra- or re-sampling.

### 8. Experimental validation results

Fig. 6 shows the confusion matrixes and ROC curves of AI classification algorithm for the seven specimen types. Correct classification of cancer areas ranges from 76.28% to 99.58%. As cancer area is the region of interest in the sampling process, we can conclude that the trained AI algorithm shows need-to-improve cancer sensitivity for rectum (76.28%) and breast(77.15%), good sensitivity for colon(83.24%) and excellent sensitivity for stomach (98.48%), esophagus (97.95%), kidney (99.51%), and lung (99.58%). From the confusion matrix and ROC curves, we can observe that the correct classification of muscle and adipose is much easier than other normal tissues. Since cancer and the normal area can have mixed areas within their boundaries and they have similar spectral characteristics which may be caused by their similarity in tissue composition, the classification of normal and cancer shows room for improvement. Therefore, extending a small distance (e.g. 2mm) between normal-cancer boundaries when sampling the specimen may help ensure the integrity of cancer area in subsequent histology examination.

**Fig. 6.**
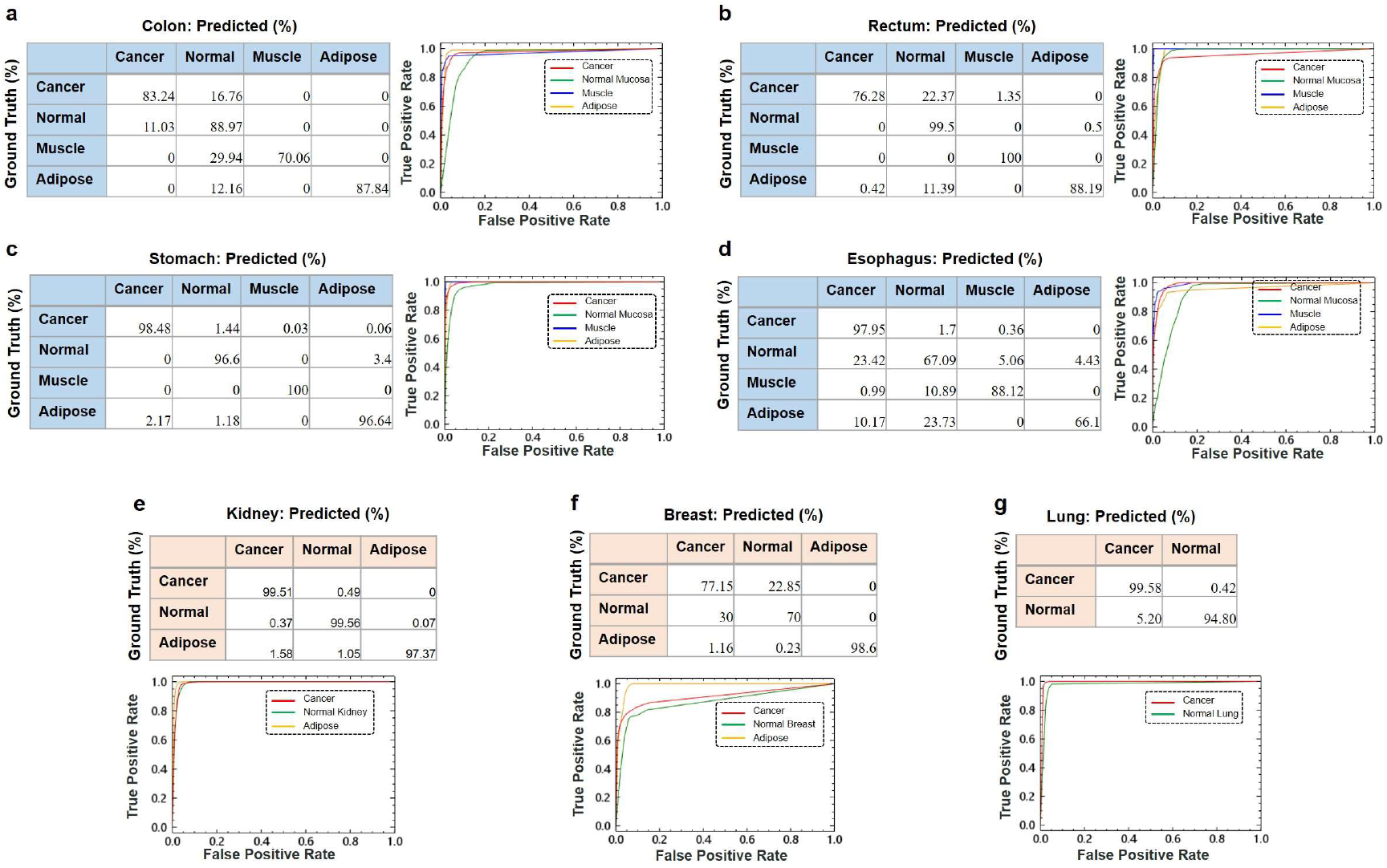
Confusion matrixes and ROC curve of the AI tissue classification algorithm for Colon (**a**), Rectum (**b**), Stomach (**c**), Esophagus (**d**), Kidney (**e**), Breast (**f**) and Lung(**g**).

To verify the ability of NIR-II HSI in identifying cancer area, we compared four approaches as mentioned in (Fig. 2d). By considering the cancer area as positive and all other areas as negative or healthy, as shown in Table. 2, for all cases the AI method demonstrates the highest sensitivity and specificity and the best consistency (sensitivity: 0.811±0.214 vs 0.880±0.151 vs 0.880±0.147 vs 0.911±0.045, specificity: 0.889± 0.158 vs 0.841±0.173 vs 0.880±0.143 vs 0.948±0.286), which proves the AI-based NIR-II HSI method is an accurate and robust technique for specimen sampling and has a significant improvement (a 14.2% increase of F1 score) beyond naked-eye observation method. Higher sensitivity means more cancer areas have been correctly identified. Low sensitivity is equivalent to missing significant cancer areas in the specimen sampling process, which is the case we want to avoid. Higher specificity means more negative or healthy areas have been correctly identified. On contrary, low specificity means significant healthy areas have been selected for histology which can cause unnecessary workload for pathology slide making and examination under the microscope. The accuracy and F1 score are both single-value metrics that help evaluate the classifier comprehensively. As we can observe from Table. 2, although for certain organs, method 1-3 can occasionally have higher sensitivity or specificity over method 4(AI), overall, the accuracy and F1 score values of method 4 remain the highest among all cases. In this manner, we can rank the tissue classification performance of the four compared methods as follows: method 4(NIR-II HSI by AI)> method 3(NIR-II color +visual) >method 2 (X-ray +visuale) ≈method 1(conventional image+visual only).

**Table.2.** Cancer identification performance of the proposed and conventional methods.

|  | Method | Sensitivity | Specificity | Accuracy | F1 score |
| --- | --- | --- | --- | --- | --- |
| All Cases | Visual | 0.811±0.214 | 0.889±0.158 | 0.871±0.126 | 0.786±0.186 |
|  | X + visual | 0.880±0.151 | 0.841±0.173 | 0.860±0.122 | 0.793±0.167 |
|  | NIR-II + visual | 0.880±0.147 | 0.880±0.143 | 0.887±0.100 | 0.824±0.185 |
|  | AI | **0.911±0.045** | **0.948±0.0286** | **0.940±0.021** | **0.898±0.037** |
| Colon | Visual | 0.781±0.189 | 0.943±0.062 | 0.896±0.068 | 0.792±0.131 |
|  | X + visual | **0.864±0.120** | 0.887±0.106 | 0.878±0.074 | 0.793±0.094 |
|  | NIR-II + visual | 0.856±0.114 | 0.922±0.079 | 0.903±0.062 | 0.826±0.083 |
|  | AI | 0.834±0.076 | **0.952±0.012** | **0.920±0.024** | **0.847±0.037** |
| Rectum | Visual | 0.787±0.194 | 0.944±0.056 | 0.913±0.048 | 0.756±0.149 |
|  | X + visual | 0.851±0.148 | 0.905±0.079 | 0.895±0.065 | 0.754±0.156 |
|  | NIR-II + visual | **0.883±0.088** | 0.927±0.069 | 0.917±0.054 | 0.809±0.111 |
|  | AI | 0.763±0.106 | **0.997±0.002** | **0.934±0.038** | **0.863±0.069** |
| Stomach | Visual | 0.714±0.248 | 0.804±0.205 | 0.789±0.130 | 0.672±0.213 |
|  | X + visual | 0.805±0.211 | 0.712±0.191 | 0.767±0.123 | 0.684±0.222 |
|  | NIR-II + visual | 0.852±0.198 | 0.739±0.181 | 0.811±0.099 | 0.736±0.218 |
|  | AI | **0.985±0.002** | **0.998±0.001** | **0.989±0.002** | **0.983±0.006** |
| Esophagus | Visual | 0.761±0.211 | **0.885±0.134** | 0.838±0.120 | 0.775±0.166 |
|  | X + visual | 0.858±0.166 | 0.833±0.140 | 0.843±0.101 | 0.807±0.127 |
|  | NIR-II + visual | 0.866±0.134 | 0.862±0.121 | 0.864±0.085 | 0.830±0.108 |
|  | AI | **0.980±0.015** | 0.850±0.105 | **0.909±0.042** | **0.871±0.078** |
| Kidney | Visual | 0.969±0.027 | 0.927±0.069 | 0.950±0.035 | 0.945±0.030 |
|  | X + visual | 0.961±0.051 | 0.912±0.086 | 0.937±0.052 | 0.928±0.051 |
|  | NIR-II + visual | 0.943±0.086 | 0.930±0.072 | 0.944±0.039 | 0.931±0.054 |
|  | AI | **0.995±0.003** | **0.985±0.001** | **0.993±0.002** | **0.990±0.003** |
| Breast | Visual | 0.758±0.302 | 0.720±0.293 | 0.728±0.240 | 0.627±0.256 |
|  | X + visual | **0.898±0.157** | 0.674±0.294 | 0.741±0.200 | 0.684±0.167 |
|  | NIR-II + visual | 0.882±0.177 | 0.795±0.223 | 0.815±0.172 | 0.742±0.170 |
|  | AI | 0.772±0.103 | **0.902±0.065** | **0.865±0.035** | **0.763±0.062** |
| Lung | Visual | 0.904±0.149 | 0.925±0.062 | 0.918±0.072 | 0.894±0.131 |
|  | X + visual | 0.944±0.104 | 0.910±0.079 | 0.925±0.063 | 0.912±0.100 |
|  | NIR-II + visual | 0.895±0.178 | 0.944±0.054 | 0.925±0.075 | 0.900±0.139 |
|  | AI | **0.995±0.003** | **0.948±0.002** | **0.971±0.002** | **0.971±0.002** |
| **Visual:** Conventional color image (Method 1). |  |  |  |  |  |
| **X + visual:** X-ray + Conventional color image (Method 2). |  |  |  |  |  |
| **NIR-II+visual:** NIR-II synthesized color image + Conventional color image (Method 3). |  |  |  |  |  |
| **AI:** AI prediction using NIR-II HSI data ((Method 4). |  |  |  |  |  |

## Discussion

This study is the first to evaluate the ability of a NIR-II HSI system to identify postoperatively resected formalin-fixed cancer tissues. Compared with conventional specimen sampling methods, NIR-II HSI provides a sensing range (900-1700 nm) beyond humans (visible 400-700 nm), showing better contrast of cancer and other tissues such as adipose, muscle, etc., thereby improving the sampling accuracy and efficiency. Statistic results show an average 14.2% (F1 score) tissue classification improvement for NIR-II HSI AI-based method over conventional naked-eye observation method. The NIR-II synthesized color image provides the pathologists with the intuitive NIR-II representation of the specimen. The AI-predicted results can be a second opinion by computationally analyzing the full spectral data. Taking the kidney cancer sample shown in Fig. 5 a3-e3 as an example, the NIR-II synthesized color image and AI-predicted results show clear cancer area, especially at the junction of cancer and the renal capsule as shown in the zoomed area, thus providing precise cancer margin to allow the sampling physician to identify the invasion degree of the renal peritoneum, enabling the physician to precisely select the site closest to the renal capsule for sampling, thus obtaining accurate histology results on the relationship between the capsule and cancer.

The advantage of the proposed system is three folds. First, it works for multiple cancer species by providing better tissue contrast and classification performance than human eye observation. This may be attributed to the more distinguishable spectral features of the different tissues in the NIR-II range. X-ray is density-dependent and only provides a monochrome channel output therefore has significant limitations in specimen sampling. In our study, the X-ray only works well for breast cancers among the multiple organs we collected. Second, the NIR-II HSI system can be used openly in the air since it has no ionizing radiation (e.g. X-ray) and is not affected by regular indoor light (350-700 nm). Therefore, the NIR-II HSI can be put right on the sampling desk with the least interruption of pathologists’ workflow. Third, the AI prediction process is automated which requires no pathologist’s observation and annotation. This technology has the potential to expedite the automation of future pathology workflow.

The proposed method still shows room for improvement. First, the SVM-based AI classification algorithm only analyzes spectral features. Hence, the spatial information, for example, the tissue texture features, and spatial continuity are not taken into calculation in the tissue classification process. 3D (spatial and spectral)-based deep learning methods take 3D data cube as input, which could be a more powerful tool in this scenario^28^. However, more training data is required for such deep learning tasks. Second, the NIR-II HSI methods provide good sampling guides by showing results on the monitor but not directly on the specimen, which can cause visual transfer errors. Therefore, augmented reality techniques can be introduced to minimize such visual transfer errors since the pathologists can focus on the specimen without turning their heads to the monitor to observe the NIR-II images and segmentation results. For example, commercial compact projectors can be integrated onto the sampling desk to project tissue boundary outlines onto the specimen for visual-transfer-free sampling navigation.

Given the benefits of NIR-II HSI in the pathology sampling process, this technique may be useful in more organ types not included in this study. The proposed method may find applications in the sampling of the cancer specimen that has undergone neoadjuvant therapy which is challenging with naked-eye observation due to the therapy response can have concentric and non-central withdrawal and the latter may have multiple scattered cancer foci in the specimen and the tumor bed is not apparent grossly^29^. Also, although our current work focused on resected formalin-fixed cancer tissues, the NIR-II HSI could be used for other pathology sampling applications, such as in vivo skin cancer biopsy, in vivo intestinal biopsy by building this system into an endoscopic form, intraoperative rapid pathology, etc. It is conceivable that the NIR-II HSI, combined with AI, could lead to many translational and clinical applications.

## Methods

### 1. Hyperspectral data reflectance correction

To reduce the influence of camera dark current and ambient light on the acquisition process, it is necessary to perform reflectance correction on the raw hyperspectral data.

As shown in the following formula,

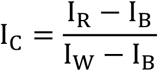

I_C_ is the reflectance corrected image. I_R_ is the raw hyperspectral image. I_B_ is the blackboard image, where the reflectance is 0%. I_W_ is the whiteboard image, where the reflectance is 99.9%.

### 2. AI segmentation algorithm

The specimen amount(92) we collected is not large enough for general deep learning tasks. However, the total spatial pixel number is huge as Table. 1 shows. As each spatial pixel correspond to a 1*256 vector (the spectral data from 900-1700 nm), we can use the each spatial pixel(or the spectral vector) as the AI input. In this study, we use the Support Vector Machine (SVM)^30^, a supervised classification method derived from statistical learning theory, for tissue classification. It separates the classes with a decision surface that maximizes the margin between the classes. While SVM is a binary classifier, it can function as a multiclass classifier by combining several binary SVM classifiers (creating a binary classifier for each possible pair of classes). We use the pairwise classification strategy for multiclass classification^31^. We also use the Radial Basis Function (RBF)^32^ as the kernel type of the SVM to map sample data to a higher dimensional space for better classification.

### 3. WSI Stitching software

We developed a WSI fragment stitching software which can perform import, rotation, mirroring, movement of multiple WSI images, and stitching of WSI fragments to restore a virtual large slice. The programming language of this WSI stitching tool is Python, and the main toolkits used are Openslide^33^ and PyQt4^34^. We make the image background transparent through image matting, which can avoid mutual occlusion between WSI fragments during the stitching process. The user starts stitching by referring to a prescanned X-ray image with the cut edge record. Once the stitching is completed and confirmed by the user, the affine transformation is recorded for the lossless reconstruction of the virtual large slide.

### 4. Co-registration between macro images and histology

Histology is the clinical gold standard for diagnosis. We use the annotation on histology as the ground truth. In our study, we use the stitched WSI to recover the virtual large slice. Co-registration between macro images (regular color images) and histology is necessary for the performance evaluation of the proposed method. We use a manual registration method^35^ to register images captured by different instruments by placing multiple (4-10) paired dots between unregistered images. The affine transformation between two modes of images is computed and recorded. Hence, the annotation on the histology (the ground truth) can be transferred onto the macro image according to the recorded affine transformation. The hyperspectral images, X-ray images, and regular color images are co-registered in the same manner. After the co-registration and annotation migration on the same image mode, the sensitivity and specificity of tissue classification can be calculated.

## Data Availability

All data produced in the present study are available upon reasonable request to the authors.

## Acknowledgements

This work is supported by the Beijing Fine Inspection Foundation, (Grant No. JJIS2021-011). This study is also supported by Tencent AI Lab, the Fourth Hospital of Hebei Medical University and the West China Hospital, Sichuan University. We thank the pathologists in the Fourth Hospital of Hebei Medical University for collecting the specimens. We thank all engineers in Tencent AI Lab for data processing and analysis of images.

## Author contributions

Y.P.L., J.H.Y, and H. B. conceived of the idea and directed the work. L.L.Z. and J.L. performed the experiments, analysed the data, wrote and edited the manuscript. M.Z., D.D.H, Z.L.J collected tissue samples and completed the collection of conventional color images, X-ray and specular images. All pathologists participated in sample labeling. H.W., C.J. and C.C assisted in data extraction of optical software. All authors wrote and revised the manuscript.

## Additional information

The study was approved by the ethics committee of the Fourth Hospital of Hebei Medical University. Written informed consent was obtained from all of participants according to the study protocols.

